# Acceptability, feasibility, quality of life and diabetes distress score outcomes: A pragmatic randomised clinical trial on continuous glucose monitoring for people with type 1 diabetes

**DOI:** 10.64898/2026.08.26.26361479

**Authors:** Elena Marbán-Castro, Lorrein Muhwava, Sarah Girdwood, Tanja Kemp, Johanè Freitas, Yvonne Kamau, Michael Otieno, Dorcas Akach, Alba Morató, Sergi Sanz, Vincent Fiechter, Berra Erkosar, Mikaela Watson, Beatrice Vetter, Cathy Haldane, Sonjelle Shilton, Paul Rheeder, Joel A Dave, Michelle Carrihill, Maria Karsas

## Abstract

**Introduction:** Continuous glucose monitoring (CGM) offers an advancement over traditional self-monitoring of blood glucose (SMBG) for people living with type 1 diabetes (T1D). However, evidence on the acceptability and feasibility of different CGM use cases in African populations remains limited.

**Methods:** This was a pragmatic three-arm, randomised controlled trial on CGM conducted among people living with T1D in three public healthcare clinics in South Africa. Participants were assigned to Arm 1 (continuous CGM), Arm 2 (periodic CGM), or Arm 3 (SMBG). Diabetes education was provided at all study visits. Feasibility was assessed by adherence to CGM use and through the Glucose Monitoring Satisfaction Survey (GMSS). Diabetes distress was measured by the Diabetes Distress Scale (DDS), health-related quality of life (HRQoL) by the EQ-5D scales, and acceptability using the Theoretical Framework of Acceptability (TFA). Surveys were collected on paper and transferred to OpenClinica. Analyses were performed in R. The trial was registered in the Clinical Trials Registry (<u>NCT05944718</u>) on July 13, 2023.

**Results:** A total of 83 participants were included in Arm 1, 85 in Arm 2, and 80 in Arm 3. CGM mean active time was 55% in Arm 1 versus 69% in Arm 2. The proportion of participants meeting the ≥70% active time threshold was higher in Arm 2 (52%) than in Arm 1 (34%). Diabetes’ distress declined across arms during the intervention period, with no significant difference between arms; distress increased slightly six months post-intervention but remained below baseline. At 6 months, glucose monitoring satisfaction was significantly higher in both CGM arms than in the SMBG arm, and satisfaction increased over time in CGM arms. Health-related quality of life remained stable across arms during the intervention period with no significant difference between arms. High acceptability was observed in both CGM arms, with higher ratings in the periodic arm.

**Conclusions:** CGM was acceptable to people living with type 1 diabetes and feasible to use in public-sector clinics in South Africa, with high acceptability under continuous and periodic use. Health-related quality of life remained stable across arms, and diabetes-related distress declined, during the intervention period, across arms. Glucose monitoring satisfaction rose significantly in both CGM arms compared to SMBG. Periodic CGM might be a promising and potentially more scalable option than continuous use for public-sector care.

*Clinical Trials registration:* NCT05944718

## Background

Diabetes mellitus (DM) represents a major public health and clinical concern worldwide. Approximately 18.7 million people in low-and middle-income countries (LMICs) are living with DM, yet it remains substantially underdiagnosed in many settings (1). In South Africa, the absence of reliable national incidence data on Type 1 diabetes mellitus (T1D) means that the burden must be inferred from regional estimates (1). The clinical benefits of continuous glucose monitoring (CGM) over traditional self-monitoring of blood glucose (SMBG) is well established (2). CGM provides continuous interstitial glucose readings, enabling detection of glycaemic patterns, including nocturnal hypoglycaemia (3). Evidence shows that CGM use is associated with reductions in HbA1c, increased time in range, and lower rates of hypoglycaemic events in people with T1D (4).

Less understood is whether CGM can be successfully implemented in public sector diabetes clinics in African countries and the conditions needed to make this feasible. Evidence from Africa remains limited, with only a small number of studies assessing CGM acceptability or feasibility (5–8). Participants with T1D in rural Malawi and their healthcare providers generally perceived CGM as acceptable when embedded within structured clinical follow-up and education (6). Qualitative interviews conducted alongside a randomised trial identified patient–provider relationships, stigma, device usability, and implications for clinical management as dominant themes shaping acceptability (6). Participants highlighted improved understanding of glucose patterns and perceived clinical benefits, although sustained uptake was contingent on ongoing education and health-system support, reflecting the operational constraints of first-level hospitals in resource-limited settings (6). Clinical feasibility data from the same programme indicated that CGM implementation in district hospitals was operationally achievable within routine diabetes services (7). CGM can be integrated into decentralized care structures despite infrastructure limitations (7).

In Europe, a multicenter randomised crossover study concluded that adults reported greater satisfaction after using CGMs, with a positive association between CGM use and patient treatment satisfaction of more than 70 % (9). Treatment satisfaction has been suggested to be an indicator of better outcomes in diabetes. Although it was anticipated that the introduction of CGM would increase anxiety and treatment burden for both parents and children, thereby reducing health-related quality of life (HR-QoL), no statistically significant differences were observed in children. Nevertheless, there was a consistent trend towards poorer scores across all HR-QoL domains, with the exception of the emotional domain (9).

A consistent gap is the lack of randomised controlled trials from South Africa evaluating implementation outcomes of CGM in public sector clinics. The ACCEDE trial was designed to fill part of this gap. As previously published, the overall objective of this trial was to assess the impact of CGM (Intervention) on glucose levels (Outcome) compared with standard of care (Comparator) in people with T1D in South Africa (Population) (10). The clinical outcomes of the ACCEDE trial, which directly address these endpoints in South Africa, are reported separately *[placeholder for clinical manuscript DOI]*.

## Methods

### Study design

This was a pragmatic three-arm, randomised controlled trial of CGM (continuous and periodic use) compared to standard of care (SMBG) in people with T1D attending public healthcare centers in South Africa. The trial included a 9-month intervention period with a 6-month follow-up. The intervention was delivered by the healthcare providers working at the sites providing diabetes care, who were also part of the research team. Care was received as of standard of care for all groups, and included diabetes education at all sites (that was not standard of care at site 01). SA Diabetes Advocacy was involved throughout the trial, from a co-creation workshop at inception through a mid-term workshop during implementation to the dissemination workshop, contributing to its design, conduct, and reporting.

### Objectives

The primary objective and first two secondary objectives are defined elsewhere *[placeholder for clinical manuscript DOI].* Secondary objectives included in this manuscript were to evaluate the acceptability and feasibility of continuous or periodic CGM device use from the perspective of the participants and caregivers.

### Location: Trial setting

The trial was conducted in three diabetes clinics across two provinces: the adult and paediatric diabetes clinic at Steve Biko Academic Hospital in Pretoria, in Gauteng (site 01), the diabetes clinic at Groote Schuur Hospital in Cape Town (site 02), and the paediatric diabetes clinic at Red Cross War Memorial Children’s Hospital in Western Cape Town (site 03).

### Population: Eligibility criteria

Eligible participants were minors (over 4 years of age) and adults with T1D whose HbA1C levels (within the past 3 months) were of ≥10%/86 mmol/mol and no record of an HbA1c < 8%/ <64 mmol/mol within the past 9 months. Participants were randomly assigned (1:1:1) to 3 arms: Arm 1) to wear the CGM continuously for 9 months, Arm 2) periodic use of CGM for 2 weeks every 3 months during that 9-month period and Arm 3) standard of care SBGM.

### Sample size calculation

The sample size was calculated to respond to the primary clinical outcome. The trial was powered to detect a 2% change in HbA1c following continuous use of a CGM device (determined by expert consultation) and assuming an effect size of 0.2. A total sample size of 246 participants (82 in each arm) was estimated to provide 80% power to detect within- and between-group differences at the 5% significance level (using repeated measures analysis of variance [ANOVA]) (10).

### Allocation concealment

Allocation concealment was ensured through centralised implementation of the randomisation sequence. A statistician from the FIND Data Science Unit generated a block randomisation list (blocks of 6 and 9) assigning participants to the study arms before enrolment began. Research assistants enrolling participants did not have access to this sequence; instead, they entered the participant’s ID into a secure electronic database (OpenClinica), which automatically returned the assigned study arm. Participants were allocated sequential unique randomisation numbers at each site, encoding group assignment according to the pre-generated schedule, thereby preventing foreknowledge of allocation and reducing risk of selection bias. As site 01 did not achieve the planned recruitment target, additional pre-generated randomisation numbers from the original allocation schedule were reassigned to site 02. This adjustment preserved the initial randomisation scheme while allowing recruitment completion without giving site investigators access to the allocation sequence.

Blinding was not done because the intervention involved wearing a visible CGM on the arm, making treatment allocation apparent to participants and healthcare providers.

### Enrolment

Enrolment took place from the 11th of September 2023 to the 6th of June 2024 across three study sites: site 01 (11-Sep-2023 to 30-Abr-2024), site 02 (25-Jan-2024 to 06-Jun-2024), and site 03 (22-Feb-2024 to 30-May-2024). People living with diabetes attending the study clinics were pre-screened. Those fulfilling eligibility criteria and not fulfilling any exclusion criteria, were invited to participate in the study. Follow up continued until the last study visit: 07-Jul-2025 for site 01 , 10-Jul-2025 for site 02 , and 10-Jul-2025 for site 03.

### Device

An intermittently scanned CGM device, Freestyle Libre 1 (FSL1) by Abbot was used. Devices were purchased from the local manufacturer (Abbott South Africa).

### Data collection

Data was collected in paper-based case report forms (CRFs) and entered into a validated electronic data capture system, OpenClinica. The following surveys were administered.

Health-related quality of life was assessed using three versions of the EQ-5D, selected according to participants’ age (11,12). The EQ-5D-3L was administered to adults (≥18 years); the EQ-5D-Y (Youth version) was completed by children and adolescents aged 8–17 years; and the EQ-5D-Proxy was completed by caregivers of children under 8 years who were unable to self-report. All versions were interviewer-administered and are well-established instruments available in English, Spanish and Portuguese (13). However, as no South African value set has been published for these instrument versions, utility scores were derived using the Zimbabwean value set for adult and proxy data and the Indonesian value set for youth data, these being the closest available geographical and socio-economic comparators (14,15). Items utilized a 0-4 Likert 286 response scale. Raw scores were transformed per published scoring instructions to obtain respective Domain, Summary and Total scores that ranged from 0-100. Higher scores reflected better functioning/HRQOL.

### Glucose Monitoring Satisfaction Survey (GMSS)

The GMSS was interviewer-administered to participants to assess satisfaction with glucose monitoring modalities, including perceived convenience, intrusiveness, trust in readings, and behavioural burden associated with device use (16). Items are scored on Likert-type response scales, with higher scores indicating greater satisfaction with monitoring practices. Domain-level scores were derived following published scoring algorithms, allowing comparison across monitoring strategies and timepoints. The instrument has been used in diabetes technology evaluations and captures both experiential and practical dimensions of glucose monitoring.

### Diabetes Distress Scale (DDS)

Diabetes-related emotional burden was measured using the DDS, a validated instrument designed to quantify regimen-related distress, interpersonal distress, physician-related distress, and emotional burden associated with living with diabetes (17,18). Items use Likert response categories reflecting frequency or intensity of distress experiences. Mean item scores were calculated according to established procedures; higher scores indicate greater diabetes-specific distress. The scale has demonstrated sensitivity to changes in diabetes management conditions, including adoption of monitoring technologies. Participants responded to the DDS themselves if they were 15 years and older. All caregivers of minors (children and adolescents under 18 years of age) responded to the caregivers DDS.

### Acceptability Questionnaire

Acceptability of CGM was assessed using a structured questionnaire informed by the Theoretical Framework of Acceptability (TFA) by Sekhon et al (19), which conceptualises acceptability as a multidimensional construct relevant during intervention implementation. The questionnaire captured seven domains: affective attitude, burden, ethicality, intervention coherence, opportunity costs, perceived effectiveness, and self-efficacy.

Acceptability and feasibility outcomes were also reported qualitatively through semi-structured interviews and focus group discussions (20).

Harms were prespecified and assessed systematically using a structured safety framework aligned with medical-device clinical investigation standards [placeholder for clinical manuscript].

### Data management and statistical analysis

Data management activities were conducted in accordance with the sponsor’s standard operating procedures (SOPs) and established principles of Good Clinical Practice (GCP). Entered data underwent routine quality control procedures, including data validation checks and query resolution with study sites to ensure completeness, consistency, and accuracy. Following completion of data cleaning and verification, the clinical database was finalized and formally locked prior to statistical analysis.

Feasibility was assessed based on adherence to the protocol in terms of CGM use. Percent participant wear time in each intervention arm was summarized by mean, median, standard deviation, minimum, maximum and quartiles. The absolute number and proportion of participants in each intervention arm depending on active CGM wear time was reported with their 95% confidence intervals (Wilson’s score method). As per the Statistical Analysis Plan (SAP), the feasibility analysis was performed on the ITT population. Missing data were not imputed, and participants without CGM data (4 in Arm 1 and 2 in Arm 2) were excluded from the feasibility analysis. Active time was defined using available CGM readings and represents a composite of sensor wear time and scan frequency during each participant’s allocated CGM wear period. In order not to lose any CGM data, participants were instructed to scan the sensor at least every 8 hours.

Health-related quality of life (pooled across the EQ-5D-3L, EQ-5D-Y, and EQ-5D-Proxy Scales) and Glucose monitoring satisfaction survey (GMMS) scores were measured at enrollment/baseline, M9, and M15, while the Diabetes Distress Score (DDS) was assessed at enrollment/baseline, M6, M9, and M15. The obtained scores were analyzed using linear mixed-effects models (LMM) including arm, visit, and their interaction as fixed effects. To adjust for potential confounding and baseline characteristics, fixed covariates were fitted to each outcome: the quality-of-life model included age, gender, and instrument format; and the GMMS and DDS models were adjusted for age and gender only. To account for the repeated measurements and the longitudinal nature of the data, a random intercept for each participant was included. Statistical significance for the fixed effects was determined via analysis of variance (ANOVA). Post-hoc pairwise comparisons were performed using estimated marginal means, with Benjamini-Hochberg’s adjustment applied within each visit to account for multiple testing across study arms. Acceptability scores at M6 and M9 were presented using descriptive statistics, describing global scores, as well as responses reported in each of the seven domains.

All analyses were performed using R software (version 4.5.0). Linear mixed-effects modelling was conducted using the lme4 and stats libraries, with post-hoc contrasts performed via the emmeans library. Data processing and table generation utilized the tidyverse and gtsummary frameworks, while all graphical visualizations were generated using the ggplot2 package.

Given the sample sizes within each arm, results in the manuscript are presented for the intention to treat (ITT) population. Analysis on the fully compliant (FC) populations are shown in Supplementary materials. A total of 14 protocol deviations were recorded: one informed-consent documentation where the witness of a visually impaired participant did not co-sign the information sheet (site 01), nine related to participants’ schedule and lack of CGM data because CGMs were not applied before the study visit or scanned as frequently as needed (site 01), three cases in which potentially identifiable participant information was shared with the sponsor (site 02) and one control-arm participant used a CGM sourced outside the study (provided by their private practitioner) for 14 days (site 03). All deviations were corrected and reported to the sponsor and if applicable, to the ethics committee. Deviations did not compromise participant safety or the scientific integrity of the study.

### Ethics

The protocol was approved by the Faculty of Health Sciences Research Ethics Committee at the University of Pretoria (330/2023) and the Human Research Ethics Committee at the University of Cape Town (HREC REF 558/2023). Written, informed consent to participate in the trial was obtained from all participants and/or their parents/caregivers.

## Results

### Trial profile and analysis populations

A total of 248 participants were enrolled, randomised and completed the baseline visit, forming the ITT population (83 participants in Arm 1, 85 in Arm 2 and 80 in Arm 3) (Figure S1). For the FC population, there were 26, 41 and 71 participants for Arms 1, 2 and 3, respectively. Sociodemographic characteristics of participants have been previously reported *[placeholder for clinical manuscript DOI]*.

**Figure 1.**
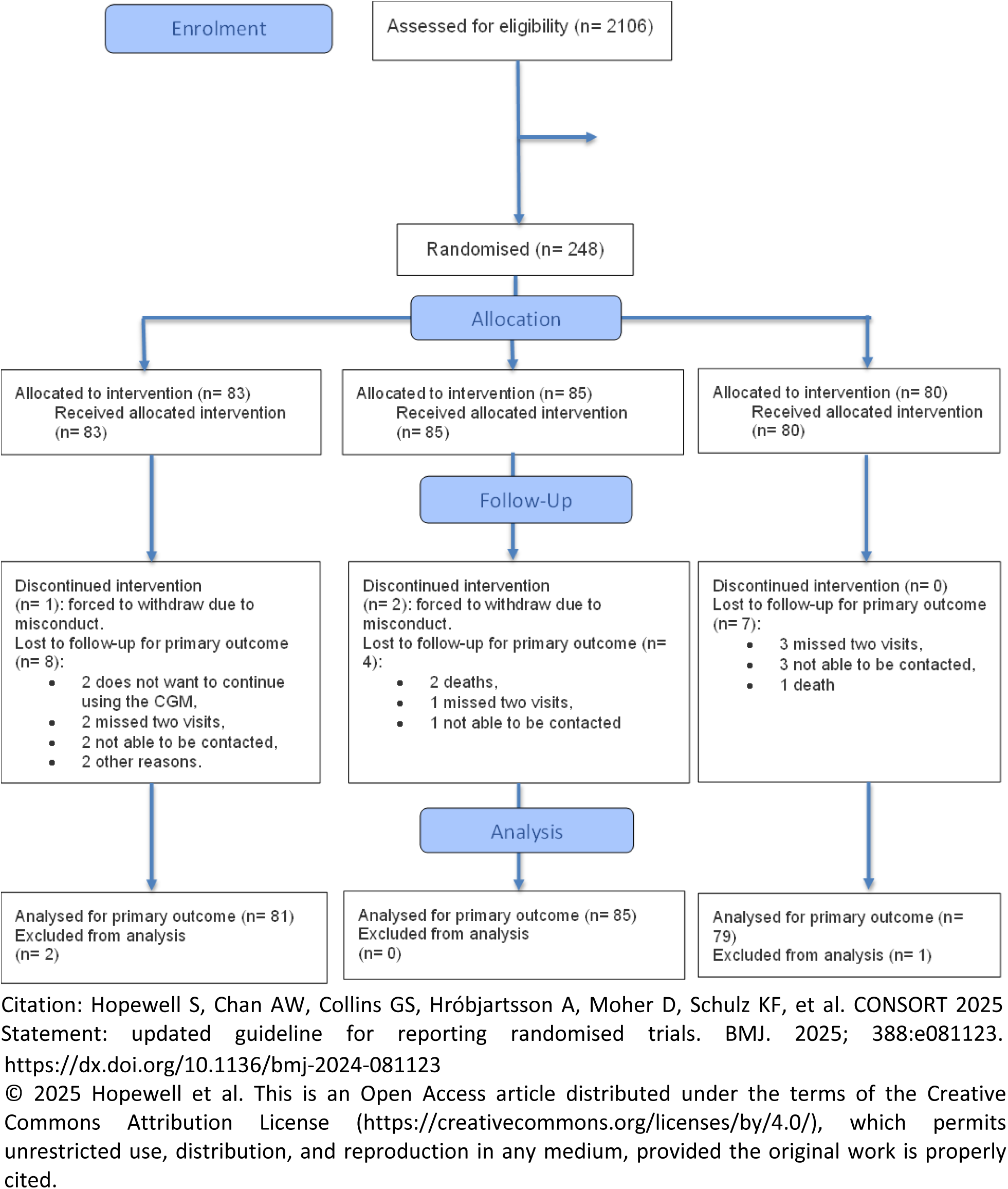
CONSORT 2025 Flow Diagram.

### Feasibility: adherence to protocol regarding CGM wear and scan for intervention arms

CGM active time differed between intervention arms. Among participants with available data at this time point, mean percentage active time reached 54.6% (SD 26.0; median 61.0; IQR 33.0–76.0; range 0–99) in Arm 1 (n=79 of 83) and 69.3% (SD 26.7; median 70.0; IQR 56.0–81.0; range 10–148) in Arm 2 (n=81 of 85), representing an estimate mean difference between groups of 14.66 (95% CI (6.44, 22.89), p=0.001). Participants achieving ≥70% total CGM active time represented 34% in Arm 1 (27/79; 95% CI 24.1% - 45.8%) and 52% in Arm 2 (42/81; 95% CI 40.5% - 63.0%). The absolute effect size was an absolute risk difference of 17.67% (95%CI (2.58%, 32.77%)), and the relative effect size was a risk ratio of 1.52 (95% CI (1.05,2.20), p = 0.024). The ≥45% adherence threshold specified for Arm 1 was met by 67% (53/79; 95% CI 55.5% - 77.0%). Arm 2 protocol adherence defined as ≥70% active in at least one monitoring period reached 75% (64/85; 95% CI 64.5% - 83.7%). As illustrated in Supplementary Figure S2 for arm 1 and Figure S3 for arm 2, only a subset of participants achieved the visit and wear-time threshold (>=70%) required for classification as FC, resulting in smaller FC sample sizes across arms: 26 (arm 1), 41 (arm 2), 71 (arm 3).

From all CGMs used by participants in Arms 1 and 2 (N=668), 179 (26.8%) CGMs were reported to have experienced an issue. The most frequently reported CGM issues were premature sensor fall-off (70/661, 10.6%), failure to scan properly (64/661, 9.7%), and sensor adhesion problems (45/661, 6.8%) (Table S1).

### Health related quality of life

Quality of life (QoL) scales were answered by 83 participants in Arm 1 (49 responded to EQ-5D-3L, 4 to EQ-5D-Proxy and 39 to EQ-5D-Y), 85 in Arm 2 (51 responded to EQ-5D-3L, 6 to EQ-5D-Proxy and 37 to EQ-5D-Y), and 80 in Arm 3 (52 responded to EQ-5D-3L, 2 to EQ-5D-Proxy and 36 to EQ-5D-Y). Quality of life reported by participants in arms 1 and 3 slightly decreased during the intervention period and remained lower at month 15 (Figure 2). Baseline mean QoL scores were 0.92 (0.12) in arm 1, 0.89 (0.16) in arm 2 and 0.93 (0.11) in arm 3. From baseline to endline (month 15), mean score changes were –0.02 (95% CI (−0.05, 0.00)) in arm 1, 0.02 (95% CI (−0.02, 0.05)) in arm 2 and –0.03 (95%CI (−0.05, 0.003)) in arm 3. The between-group difference mean change from baseline to endline was not statistically significant: the estimated mean difference between arm 1 and arm 2 was –0.02 (95% CI (−0.09, 0.05), p=0.407) and between arm 1 and arm 3 was 0.07 (95% CI (−0.02, 0.15), p = 0.086). Responses per survey by age group are shown in Figure S4. For the FC population (n = 27 in arm 1, n=48 in arm 2 and n=81 in arm 3), results were consistent with the ITT population for Arms 1 and Arm 3, b whereas Arm 2 participants showed a mean QoL increase from baseline to endline of 0.03 (95%CI (−0.01, 0.07)) (Figure S4, Supplementary).

**Figure 2.**
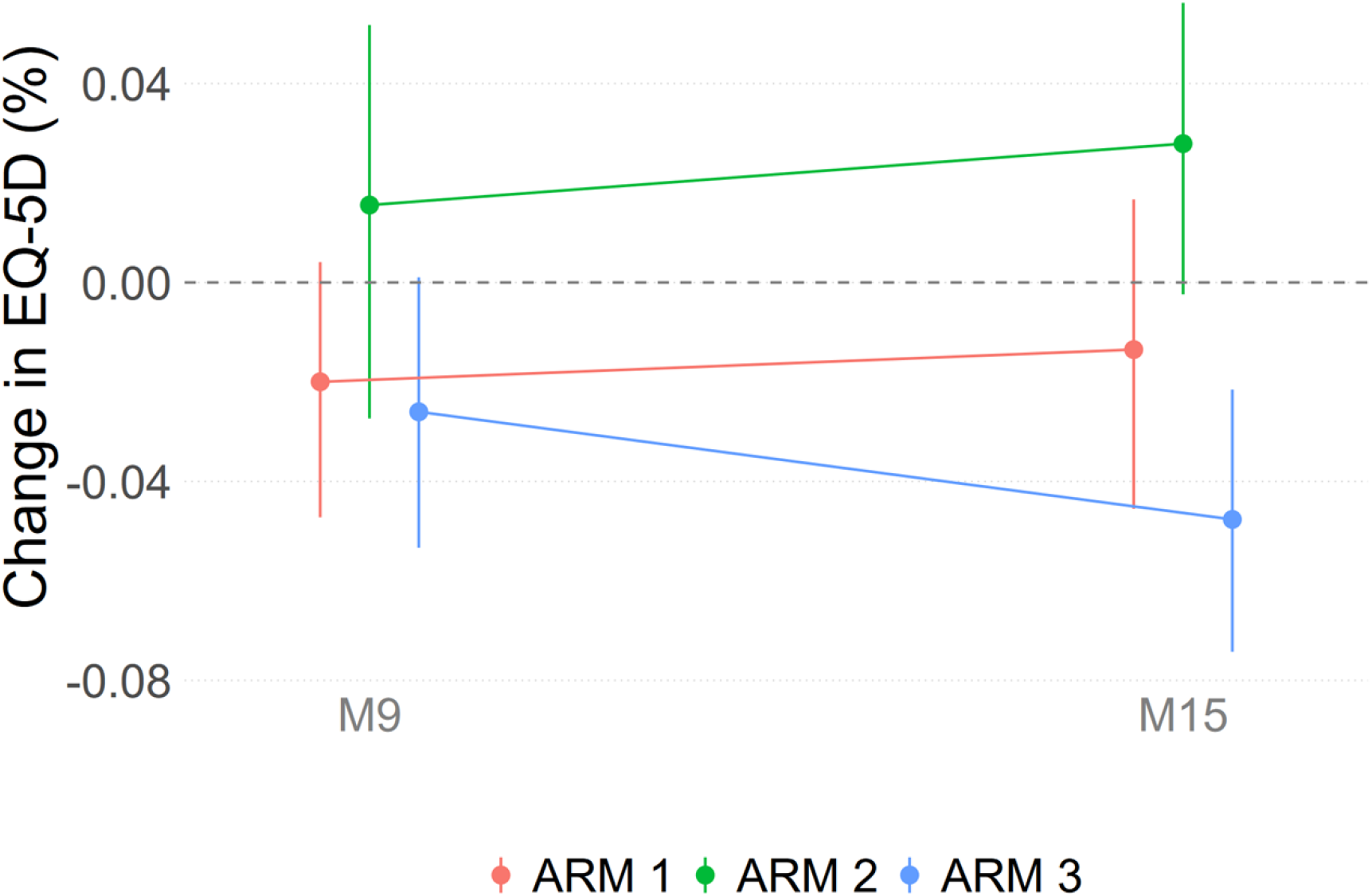
Change in health-related quality of life levels (%) from baseline to months 9 (end of intervention) and 15 (end of follow up) across study arms for the ITT population. Adjusted Change in EQ-5D from D1: Estimated mean change from baseline derived from the adjusted ANCOVA model. M: month.

### Diabetes distress scale

The DDS was collected at four timepoints, separately for both participants with at least one completed questionnaire (n = 55 in arm 1, n = 58 in arm 2 and n = 58 in arm 3) and caregivers (n = 46 in arm 1, n = 48 in arm 2 and n = 40 in arm 3). Baseline participant DDS were 80.0 (28.8) in arm 1, 78.1 (28.2) in arm 2 and 75.6 (26.1) in arm 3. Participant-reported diabetes distress declined from baseline through months 6 (mean change: -10.66; 95%CI (−15.7, -5.62)) and 9 (mean change: -13.66; 95% CI (−18.7, -8.59)) across all arms, with a rebound six months after follow up (Table 1, Figure 2A, Table S2). However, between-arm differences were not statistically significant at any visit: at month 9, the estimated mean difference was 0.03 (95%CI (−0.42, 0.47); p=0.881) for arm 1 vs arm 2, and 0.07 (95%CI (−0.37, 0.52); p = 0.881) for arm 1 vs arm 3 (Table S3). Among specific domains, eating distress and powerlessness showed the greatest reduction from baseline to month 9 (mean reduction: 1.97; 95% CI (1.14, 2.82)) (Table 1, Figure S5).

**Table 1.** Participants’ diabetes distress scales and subdomains for the ITT population during the 9 months intervention period and subsequent 6 months follow up.

|  | Week 1 |  |  | Month 6 |  |  | Month 9 |  |  | Month 15 |  |  |
| --- | --- | --- | --- | --- | --- | --- | --- | --- | --- | --- | --- | --- |
|  | ARM 1<br>N= 48 | ARM 2<br>N= 51 | ARM 3<br>N= 51 | ARM 1<br>N= 45 | ARM 2<br>N= 50 | ARM 3<br>N= 51 | ARM 1<br>N= 48 | ARM 2<br>N= 49 | ARM 3<br>N= 49 | ARM 1<br>N= 49 | ARM 2<br>N= 52 | ARM 3<br>N= 53 |
| **Total Diabetes Distress Score** |  |  |  |  |  |  |  |  |  |  |  |  |
| Mean (SD) | 80.0 (28.8) | 78.1 (28.2) | 75.6 (26.1) | 66.7 (22.2) | 67.8 (24.1) | 66.7 (27.6) | 64.9 (26.1) | 65.4 (23.0) | 62.2 (22.9) | 74.2 (28.1) | 70.0 (27.0) | 67.5 (26.4) |
| Median (Q1, Q3) | 76.5 (61.5, 97.0) | 72.0 (55.0, 99.0) | 73.0 (56.0, 95.0) | 68.0 (50.0, 80.0) | 64.5 (48.0, 83.0) | 60.0 (44.0, 81.0) | 59.5 (44.0, 82.0) | 64.0 (45.0, 82.0) | 57.0 (46.0, 72.0) | 69.0 (53.0, 93.0) | 66.0 (46.5, 88.0) | 58.0 (46.0, 83.0) |
| **Subscales** |  |  |  |  |  |  |  |  |  |  |  |  |
| **Powerlessness** |  |  |  |  |  |  |  |  |  |  |  |  |
| Mean (SD) | 17.3 (6.4) | 16.4 (6.6) | 16.5 (6.4) | 14.5 (5.8) | 14.9 (5.9) | 14.5 (6.5) | 13.4 (6.1) | 13.9 (5.7) | 13.5 (6.0) | 16.2 (7.2) | 14.9 (6.0) | 14.5 (6.3) |
| Median (Q1, Q3) | 17.5 (13.5, 22.0) | 15.0 (11.0, 22.0) | 16.0 (12.0, 22.0) | 14.0 (10.0, 18.0) | 14.0 (9.0, 19.0) | 14.0 (10.0, 19.0) | 12.5 (7.5, 18.5) | 13.0 (10.0, 17.0) | 12.0 (8.0, 17.0) | 16.0 (10.0, 23.0) | 14.5 (9.5, 18.5) | 13.0 (10.0, 19.0) |
| **Management Distress** |  |  |  |  |  |  |  |  |  |  |  |  |
| Mean (SD) | 11.8 (6.3) | 11.8 (4.8) | 11.5 (4.8) | 9.7 (4.3) | 10.0 (4.4) | 10.8 (4.9) | 9.9 (5.2) | 9.5 (4.4) | 9.4 (3.4) | 11.9 (5.7) | 10.7 (4.9) | 10.5 (4.7) |
| Median (Q1, Q3) | 9.5 (7.5, 16.5) | 11.0 (8.0, 16.0) | 11.0 (7.0, 15.0) | 10.0 (6.0, 13.0) | 8.5 (6.0, 13.0) | 10.0 (7.0, 14.0) | 9.5 (5.0, 13.0) | 8.0 (6.0, 12.0) | 9.0 (7.0, 11.0) | 12.0 (6.0, 16.0) | 10.5 (6.0, 14.5) | 10.0 (7.0, 14.0) |
| **Hypoglycemia Distress** |  |  |  |  |  |  |  |  |  |  |  |  |
| Mean (SD) | 11.0 (5.2) | 11.9 (6.3) | 10.8 (5.4) | 9.3 (3.8) | 10.2 (5.1) | 9.3 (5.0) | 8.8 (4.4) | 9.7 (4.3) | 8.6 (4.4) | 10.4 (5.3) | 9.8 (5.5) | 9.2 (4.2) |
| Median (Q1, Q3) | 10.5 (7.0, 14.5) | 11.0 (6.0, 16.0) | 9.0 (7.0, 14.0) | 9.0 (7.0, 12.0) | 9.0 (6.0, 13.0) | 8.0 (6.0, 12.0) | 8.0 (6.0, 11.0) | 9.0 (7.0, 12.0) | 7.0 (6.0, 11.0) | 9.0 (6.0, 14.0) | 8.0 (6.0, 13.0) | 8.0 (6.0, 12.0) |
| **Negative Social Perceptions** |  |  |  |  |  |  |  |  |  |  |  |  |
| Mean (SD) | 10.3 (5.5) | 9.0 (5.2) | 9.0 (6.1) | 8.5 (4.6) | 7.5 (4.1) | 8.2 (5.5) | 8.6 (4.5) | 8.0 (4.6) | 8.2 (5.1) | 9.3 (5.2) | 8.3 (4.8) | 8.6 (5.1) |
| Median<br>(Q1, Q3) | 9.0<br>(6.0, 13.5) | 7.0<br>(4.0, 12.0) | 6.0<br>(5.0, 11.0) | 7.0<br>(4.0, 11.0) | 6.0<br>(4.0, 10.0) | 5.0<br>(4.0, 11.0) | 7.0<br>(5.0, 11.5) | 7.0<br>(4.0, 9.0) | 6.0<br>(4.0, 10.0) | 8.0<br>(5.0, 12.0) | 7.0<br>(4.0, 11.0) | 7.0<br>(4.0, 11.0) |
| **Eating Distress** |  |  |  |  |  |  |  |  |  |  |  |  |
| Mean<br>(SD) | 10.3<br>(4.3) | 10.2<br>(4.3) | 9.4<br>(3.6) | 8.9<br>(4.4) | 8.7<br>(3.8) | 8.4<br>(3.6) | 8.1<br>(4.0) | 8.2<br>(3.5) | 7.5<br>(3.1) | 9.2<br>(4.2) | 9.3<br>(4.2) | 8.6<br>(3.6) |
| Median<br>(Q1, Q3) | 10.5<br>(7.0, 13.5) | 10.0<br>(7.0, 15.0) | 9.0<br>(7.0, 12.0) | 8.0<br>(6.0, 12.0) | 8.0<br>(6.0, 11.0) | 8.0<br>(5.0, 11.0) | 7.0<br>(5.0, 11.0) | 7.0<br>(6.0, 10.0) | 7.0<br>(5.0, 9.0) | 9.0<br>(6.0, 12.0) | 9.0<br>(5.5, 12.0) | 8.0<br>(6.0, 11.0) |
| **Physician Distress** |  |  |  |  |  |  |  |  |  |  |  |  |
| Mean<br>(SD) | 7.5<br>(4.1) | 7.5<br>(4.1) | 7.1<br>(4.0) | 6.5<br>(3.3) | 6.6<br>(3.8) | 5.9<br>(2.9) | 6.7<br>(4.6) | 6.1<br>(2.7) | 5.9<br>(2.9) | 7.0<br>(3.5) | 6.9<br>(3.7) | 7.0<br>(4.3) |
| Median<br>(Q1, Q3) | 7.0<br>(4.0, 9.0) | 7.0<br>(4.0, 8.0) | 5.0<br>(4.0, 8.0) | 5.0<br>(4.0, 8.0) | 5.0<br>(4.0, 7.0) | 5.0<br>(4.0, 6.0) | 5.0<br>(4.0, 7.0) | 5.0<br>(4.0, 7.0) | 4.0<br>(4.0, 6.0) | 6.0<br>(4.0, 9.0) | 6.0<br>(4.0, 9.0) | 5.0<br>(4.0, 8.0) |
| **Friend/Family Distress** |  |  |  |  |  |  |  |  |  |  |  |  |
| Mean<br>(SD) | 11.9<br>(5.6) | 11.4<br>(6.1) | 11.3<br>(5.3) | 9.2<br>(4.4) | 9.9<br>(5.0) | 9.7<br>(5.9) | 9.4<br>(4.5) | 10.0<br>(4.3) | 9.1<br>(4.8) | 10.2<br>(5.4) | 10.1<br>(5.2) | 9.3<br>(4.8) |
| Median<br>(Q1, Q3) | 10.5<br>(7.5, 17.0) | 10.0<br>(6.0, 16.0) | 11.0<br>(7.0, 15.0) | 8.0<br>(6.0, 12.0) | 9.5<br>(6.0, 12.0) | 7.0<br>(5.0, 15.0) | 9.0<br>(5.0, 12.5) | 11.0<br>(6.0, 12.0) | 8.0<br>(5.0, 11.0) | 10.0<br>(5.0, 14.0) | 9.0<br>(6.0, 13.0) | 8.0<br>(6.0, 12.0) |

**Figure 2.**
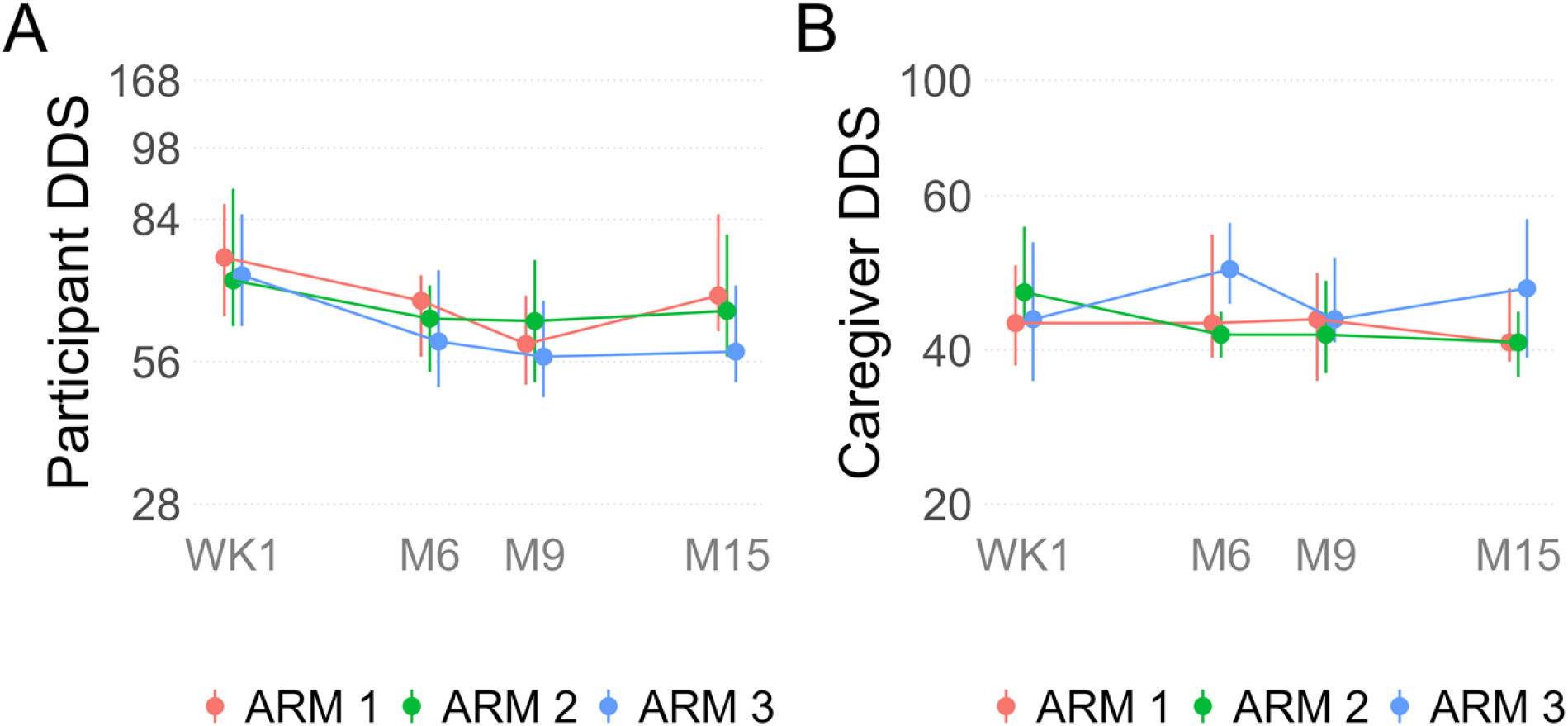
Diabetes distress scores on diabetes distress at baseline, month 6, month 9 (end of the intervention) and at month 15 (end of follow up) for the ITT population. A) Participant’s DDS, B) Caregivers DDS. The ranges for the scales were different for the participant and caregiver scores.

For caregivers, overall DDS did not differ significantly across visits or between arms. However, a significant arm-by-visit interaction (p = 0.027) was observed, reflecting a greater decline in distress in Arm 2 relative to Arm 1 at month 6 (between-group difference in mean change: -7.3; 95% CI (−14.0, -1.0)) and month 15 (between-group difference in mean change: -7.0; 95% CI (−13.0, -0.71)). At month 9, the difference between arm 2 and arm 1 did not reach statistical significance (−6.0; 95%CI (−12.0, 0.21)) (Figure 2B; Table S2; Table S3).

In the FC population (n= 16 in arm 1, n = 27 in arm 2 and n = 51 in arm 3), participants’ diabetes distress patterns remained consistent with the primary ITT findings (Supplementary Figure S6).

### Glucose monitoring satisfaction

For the ITT population with at least one completed questionnaire (n = 50 in arm 1, n = 54 in arm 2 and n = 54 in arm 3), GMSS was similar for all participants at baseline: 2.98 (0.35) in arm 1, 3.02 (0.38) in arm 2 and 3.02 (0.34) in arm 3. After having used the CGMs for six months, satisfaction with their glucose monitoring increased, statistically, for those in Arm 1 (3.51 (0.29)) and Arm 2 (3.60 (0.22)) relative to Arm 3 (2.96 (0.32)) (Figure 3 and Table S4). The estimated effect size between Arm1 and Arm 3 was 0.54 (95% CI (0.39, 0.70); p < 0.001), and between Arm 2 and Arm 3 was 0.63 (95% CI (0.48, 0.79); p < 0.001). There was no significant difference between Arm 1 and Arm 2 –0.09 (95% CI (−0.25, 0.07), p = 0.164). At month 15, end of the intervention period (where all participants have used SMBG during six months), satisfaction decreased for those in Arm 1 (3.09 (0.40)) and Arm 2 (3.28 (0.42)) but remained higher than those in Arm 3 (2.97 (0.35)) (Figure 3). At this time point, there was statistically significant differences across all arms. The estimated effect size between Arm 1 and Arm 2 was -0.19 (95% CI (−0.34, -0.03); p = 0.007). Satisfaction remained significantly higher in Arm 1 compared to Arm 3 0.13 (95% CI (−0.02, 0.29); p = 0.041), and in Arm 2 compared to Arm 3 0.32 (95% CI (0.16, 0.48); p < 0.001) (Table S4).

**Figure 3.**
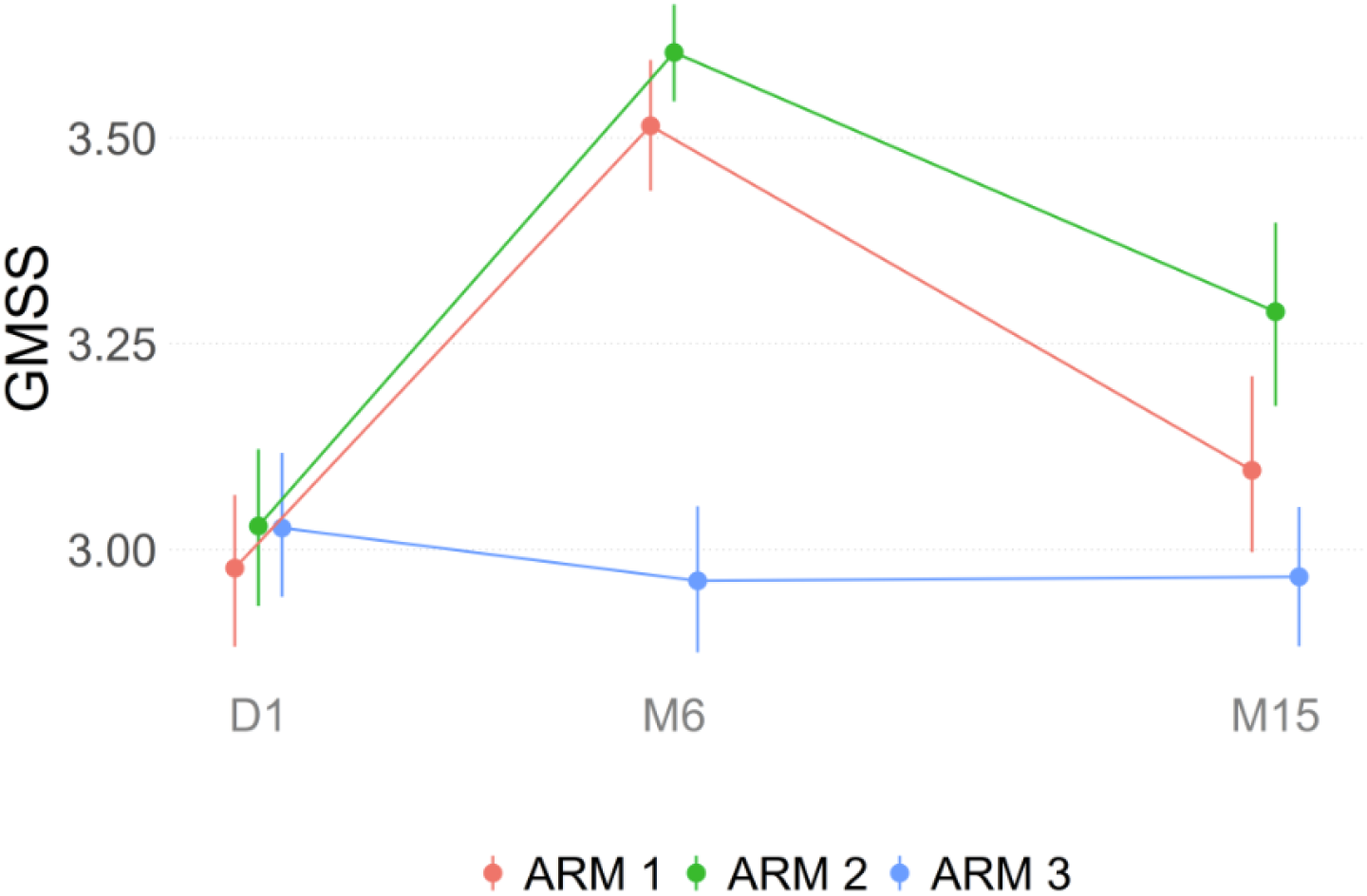
Observed GMSS levels (%) over time across the trial arms. The solid points represent the sample mean at each visit, and the vertical error bars denote the 95% confidence interval for the mean calculated using bootstrapping. WK: week; M: month.

### CGM acceptability

General acceptability was high in both CGM arms (>99% at month 6 for both arms, 93% for the continuous arm at month 15 and 97% for the period arm) (Figure 4). Acceptability was consistently higher in the periodic CGM arm compared to the continuous arm. More than 90% of participants in both arms reported that they felt confident using CGM. More than 65% of participants in both arms reported that using the CGM helped them improve their HbA1c and that they liked using the CGM. Similarly, acceptability was high for the FC population (Figure S7).

**Figure 4.**
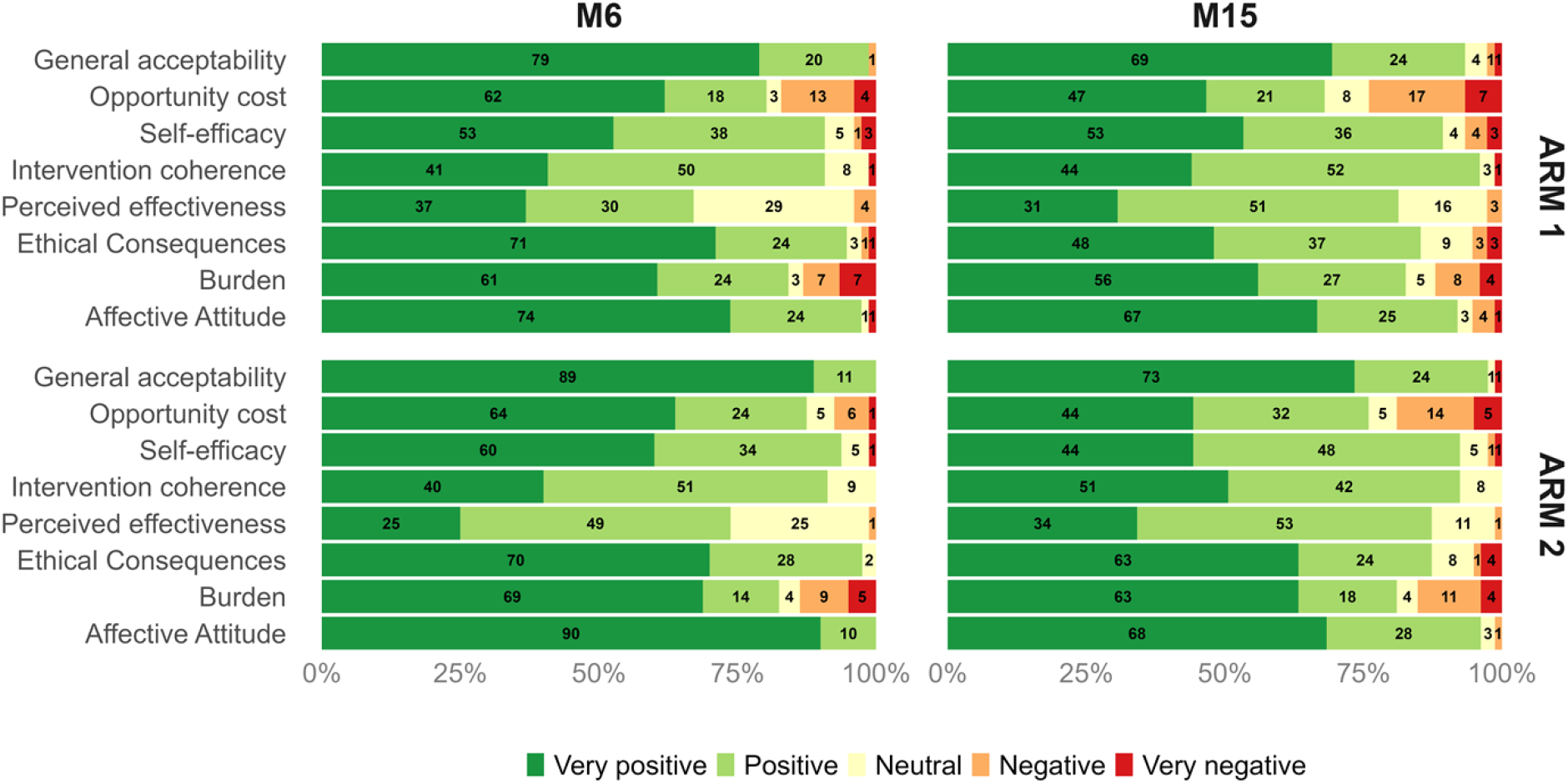
Acceptability levels (%) over time in the intervention arms for the ITT population. M: month

### Harms

Detailed hospitalisations are reported in a separate manuscript. During the 9-month intervention period there were 76 hospitalizations in Arm 1, 78 in Arm 2 and 72 in Arm 3.

CONSORT checklist is reported in Supplementary information as Figure S8.

## Discussion

To our knowledge, this is the first randomised clinical trial to report implementation outcomes of CGM use in people living with T1D using CGMs, continuously or periodically, compared to the standard of care in Southern Africa. Overall, CGM was feasible and acceptable across both use cases. CGM adherence differed between the two intervention arms. Mean active time was lower in the continuous CGM arm (55%) than in the periodic CGM arm (69%), and the proportion of participants meeting the ≥70% threshold was correspondingly higher in Arm 2 (52% vs. 34%). The continuous CGM arm required sustained, uninterrupted wear over 9 months, which could be understood as a demanding intervention. More than two thirds of participants in Arm 1 met the lower, arm-specific threshold of ≥45% active time, suggesting meaningful engagement even where optimal active time was not achieved. Low adherence to CGM might be a combination of device adhesion issues, lack of awareness or education of the benefits of frequent scanning or other issues. In our study, diabetes distress declined across all three arms during the nine-month intervention period. This finding is encouraging, but cannot be attributed to CGM alone, given that structured diabetes education was provided to participants in all study arms at every study visit.. The domains showing the greatest decline were powerlessness and eating distress, which may reflect the relevance of glucose self-management confidence in this population. Satisfaction with glucose monitoring increased in both CGM arms over the course of the intervention and declined after its end, when participants returned to SMBG. This pattern was not observed in the SMBG arm, where satisfaction remained stable throughout. This trajectory is consistent with a direct effect of the CGM on the perceived value of CGM. The return towards baseline at follow-up reinforces the findings from the acceptability analysis; the benefits participants experienced could be related to active CGM use. Acceptability was high across both CGM arms, with more than 97% of participants reported liking or strongly liking CGM. These levels of acceptability are particularly notable given the population: people with elevated baseline HbA1c attending public sector clinics, most of whom had no prior experience with CGM. Health-related quality of life utility values remained broadly stable across all arms during the intervention period, with no significant differences between CGM and SMBG arms overall. The absence of large HRQoL differences between arms was not unexpected in a pragmatic trial of this duration. The generalisability of the study findings is limited by the restricted geographic scope, relatively small sample size, and the conduct of the study within externally supported programmes, which may differ from routine healthcare settings across Africa. Such constraints complicate extrapolation to national diabetes programmes, particularly where financing, workforce capacity, and device procurement pathways differ substantially across settings.

Achieving satisfactory adhesion of the CGM to the skin, through supplemental products such as liquid adhesive, adhesive wipes, transparent dressings, tape, and wraps, can greatly improve successful CGM wear and acceptance in children (21). The choice of adhesive products should consider costs, as well as the impact of seasonal temperature variations and activities, such as sports and swimming (21). A study in Rwanda, with 50 people living with T1D, reported that participants used the CGMs more than 80% of time throughout the study period (22). However, that study included only adult participants, from 21 years onwards (22). The same cohort reported sustained clinical benefits of CGMs, reduced HbA1c and time in range, over a period of 24 months (23). High diabetes distress has been previously associated with participants presenting with hyperglycaemia (24). A study conducted in Germany showed high satisfaction with CGMs, 98% of the CGM users reported better well-being with CGM compared with previous SMBG use (25). However, diabetes distress was not associated with different glucose monitoring (SMBG 5.6 vs. iscCGM 6.2 vs. rtCGM 6.5; *P* = .386) (25). The Malawi studies, which are among the only comparable data points in Africa, similarly found high perceived acceptability when CGM was embedded in structured follow-up, but also showed that acceptability did not by itself ensure sustainability in the absence of reliable supply chains and clinical support (6,7). Others have shown acceptability of CGM use by youth people living with T1D together with a physical activity intervention (26). Physical activity should be reinforced during diabetes education sessions. The creation of groups of people living with T1D, using CGMs to share their experiences has also been shown to be acceptable, feasible, and a way to engage participants, contributing to health behaviours over time (27).

Several limitations should be noted. Participants were recruited based on elevated baseline HbA1c, reflecting challenges in glycaemic control. While this was intentional, the aim was to target those most likely to benefit from the intervention, this also means the sample may include individuals with pre-existing adherence difficulties, irregular attendance, or lower motivation for self-management, who may be less inclined to engage with CGM regardless of the intervention. The pragmatic nature of the trial, including broad inclusion criteria, means the sample also included individuals with food insecurity, psychosocial distress, and mental health concerns. These factors likely contributed to variability in adherence, higher rates of hospitalisation, and difficulty sustaining engagement, particularly among adolescents, a group for whom the emotional and social demands of this developmental stage intersect with the burden of T1D self-management in ways that generic diabetes education may not adequately address. Participants also had more frequent contact with clinical staff than they would in routine care, which may limit generalisability. Additionally, the follow-up period may have been too short to capture meaningful change in utility or quality of life; generic preference-based measures such as the EQ-5D are known to have limited responsiveness and pronounced ceiling effects, which can obscure genuine change over brief intervals, including in populations with T2D and elevated HbA1c (28,29). While validated instruments were used, their ability to fully capture quality of life and diabetes distress in this setting is uncertain. Although the EQ-5D is widely applied across LMICs, its cross-cultural transferability typically requires local adaptation, and psychometric evidence in these contexts remains limited (30). The absence of significant findings may therefore reflect the small sample size, the short observation window, and/or the instruments’ limited sensitivity in this population, rather than a true absence of effect.

Future studies should explore the relative contributions of CGMs and the education component and whether distress reductions can be sustained beyond the active intervention period. Our findings might be relevant to national discussions on how to integrate CGM into public sector T1D care in South Africa and the region.

## Conclusions

CGM was acceptable to people living with type 1 diabetes and feasible to use in public-sector clinics in South Africa, with high acceptability under both continuous and periodic use. Health-related quality of life remained broadly stable across all arms, with no significant differences between the CGM and SMBG groups, and diabetes-related distress declined across all three arms during the intervention period, a reduction that cannot be attributed to CGM specifically. Notably, acceptability under periodic CGM was comparable to continuous use while adherence was higher, and monitoring satisfaction rose in both CGM arms relative to SMBG. Combined with its lower device cost, this positions periodic CGM as a promising and potentially more scalable option than continuous use for public-sector care. Realising this potential will require sustained patient support and continued strengthening of diabetes education to translate acceptability into durable changes in health behaviour.

## Data Availability

De-identified individual participant-level?data?is available at https://zenodo.org/records/21531500. ?Study protocol?was already published at:?https://link.springer.com/article/10.1186/s13063-024-08132-7.?

## Declarations

### Authors contributions

Conceptualization: EM-C, LM, BE, BV, CH, SSh, PR, JAD, MC, MK.

Data curation: MO, DA, AM, SS.

Formal analysis: AM, SS.

Funding acquisition: BV, SSh, CH.

Investigation: EM-C, LM, SG, TK, JF, PR, JAD, MC, MK.

Methodology: E-MC, LM, SG, TK, JF, PR, JAD, MC, MK.

Project administration: EM-C, LM, VF, BV, CH, SSh.

Software: MO, DA, AM, SS.

Supervision: EM-C, LM, BV, CH, SSh, PR, JAD, MC, MK.

Visualization: AM, SS.

Writing – original draft: EM-C.

Writing – review & editing: EM-C, LM, SG, TK, JF, YK, MO, DA, AM, SS, VF, BE, MW, BV, CH, SSh, PR, JAD, MC, MK

### Consent for publication

No participant details, images, or videos are included in this trial. This section is therefore not applicable.

### Availability of data and materials

De-identified individual participant-level data is available at the following link https://zenodo.org/records/21531500. Study protocol was already published (24).

### Competing interest

Abbot sponsored registration (Prof Karsas and Prof Rheeder) for the SEMDSA (Society for Endocrinoloy, Metabolism and Diabetes of South Africa) congress in Durban 2025. Prof JA Dave has received honoraria from Abbot for continuing medical education activities on continuous glucose monitoring.

### Funding

This trial was sponsored by FIND, with a grant from The Leona M. and Harry B. Helmsley Charitable Trust. The grant code is HCT-NCDS01. As per the funding contract between The Leona M. and Harry B. Helmsley Charitable Trust and FIND, the funder had reviewed the manuscript before submission with a focus on wording relating to the funding source. The funders had no role in study design, data collection and analysis, decision to publish, or preparation of the manuscript.

## Acknowledgements

We thank all study participants and their families. We would also like to acknowledge all staff that were key to study completion: Razana Allie, Cleon Lekabe, Mary Jane Van Zyl, Mpho Simila, Prof Ian Ross, Dr William Toet, Dr Sophie Davies-Van Es, Dr Melanie Moyo, Dr Maleeka Abrahams-Kahaar, Dr Hanadi Alganeeb, Buyelwa Majikela-Dlangamandla, Neil Meiring, Tyamkazi Nqekeza, Lisa van Wyk, Dr Wayne S. Rajah, Dr Amith Ramcharan, Dr Lindsay Rajah, Dr Deepika Goolab, Laura Symmonds, Lindi Gqamana, Santie Horn, Sergi Sanz, Joseph Ndungu, Ntombi Sigwebela, Dayo Adetifa, Emilie Alirol, and Olga Denisiuk.

## Abbreviations

CGM: continuous glucose monitoring
CI: confidence interval
DDS: Diabetes Distress Score
FGD: Focus Group Discussion
GMSS: Glucose Monitoring Satisfaction Survey
HRQoL: Health Related Quality of Life
HbA1c: glycated haemoglobin
IRB: Institutional Review Board
HCP: Health care provider
LMIC: low- and middle-income countries
NHLS: National Health Laboratory Service
PRECIS: PRagmatic Explanatory Continuum Indicator Summary
SAE: serious adverse event
T1D: type 1 diabetes

## Notes

### Clinical Trial

https://clinicaltrials.gov/study/NCT05944718

### Clinical Protocols

https://pubmed.ncbi.nlm.nih.gov/38773658/

### Author Declarations

The protocol was approved by the Faculty of Health Sciences Research Ethics Committee at the University of Pretoria (330/2023) and the Human Research Ethics Committee at the University of Cape Town (HREC REF 558/2023).

